# Protein Quantitative Trait Loci Identify Genetically Regulated Immune Proteins Associated with Tuberculosis Progression in People with HIV

**DOI:** 10.64898/2026.03.27.26349487

**Authors:** Simon Boutry, Marius Zeeb, Cédric Dollé, Cornelia Staehelin, Alexandra Calmy, Matthias Cavassini, Lucas Boeck, Luigia Elzi, Patrick Schmid, Irene A Abela, Fergal J Duffy, Jacques Fellay, Johannes Nemeth

**Author notes:** Corresponding author: Johannes Nemeth. These authors contributed equally as co-senior authors (Jacques Fellay, Johannes Nemeth).

## Abstract

Host genetics alone poorly predicts tuberculosis (TB) progression, particularly in people with HIV (PWH). We performed exploratory cis-protein quantitative trait locus (pQTL) mapping in pre-diagnostic plasma from 60 PWH who later developed active TB and 194 matched non-progressing controls in the Swiss HIV Cohort Study. Stratified analyses identified 26 pQTLs linked to 12 proteins among progressors and 107 pQTLs linked to 14 proteins among controls, with no overlapping proteins between groups. The progressor-associated protein set, including HLA-C, C4B, and CHIT1, was enriched for immune-related Gene Ontology terms. These exploratory, genetically anchored candidates warrant replication and functional validation before consideration as TB biomarkers.

## BACKGROUND

Tuberculosis (TB) remains the world’s leading infectious cause of death: an estimated 10.7 million people fell ill and 1.23 million died in 2024 [1]. HIV coinfection markedly raises TB risk, and people with HIV (PWH) accounted for 5.8% of new TB cases that year [1,2]. Genome-wide association studies have identified only a handful of TB susceptibility loci, with modest effect sizes; a recent multi-ancestry meta-analysis of over 33,000 cases and controls found that common variants explain just 26.3% of TB heritability [3]. Non-coding variants of unclear function, heterogeneous phenotyping, and strong environmental confounding limit what genomics alone can reveal about TB pathogenesis, a problem compounded in HIV coinfection, where CD4-dependent immune suppression reshapes the host response to *Mycobacterium tuberculosis* (MTB) [2,4].

Protein quantitative trait loci (pQTLs), germline variants that regulate circulating protein levels, offer a complementary approach. Because the underlying germline variants are fixed at conception, pQTLs can anchor variation in protein abundance to inherited genetic control, complementing differential proteomic signatures that cannot separate causal, consequential, and environmentally driven changes. Large biobank studies have mapped thousands of pQTLs and enabled causal inference through Mendelian randomization [5,6]. Applying this approach to a cohort with archived pre-diagnostic samples could reveal genetically regulated proteins relevant to TB progression before disease onset.

We present, to our knowledge, the first pQTL analysis in PWH who progressed to active TB, using the Swiss HIV Cohort Study (SHCS) [7], which retains longitudinal plasma samples collected before TB diagnosis. We conducted exploratory, stratified cis-pQTL mapping in matched progressors and non-progressors to generate hypotheses about immune pathways associated with later TB progression.

## METHODS

This study used the SHCS, a nationwide, prospective, multicenter cohort enrolling PWH since 1988, with longitudinal plasma samples and clinical data from over 21,800 participants [7]. The SHCS was approved by the participating institutions’ ethics committees (BASEC-Nr. 2023-02080); all participants gave written informed consent.

We identified 91 SHCS participants with active TB (microbiologically confirmed or clinically defined) and an archived plasma sample ≥6 months before diagnosis. For each, we selected the sample closest to diagnosis within a 6-12-month window [8], yielding 60 TB progressors. Controls (initially n=293) without TB history or preventive therapy, were matched by a weighted algorithm [8] on age, sex, BMI, ethnicity, CD4 count, HIV viral load, and transmission category, had samples collected within ±90 days of the matched case, and had ≥5 years follow-up without TB; 194 controls entered the pQTL analysis (Figure 1).

**Figure 1.**
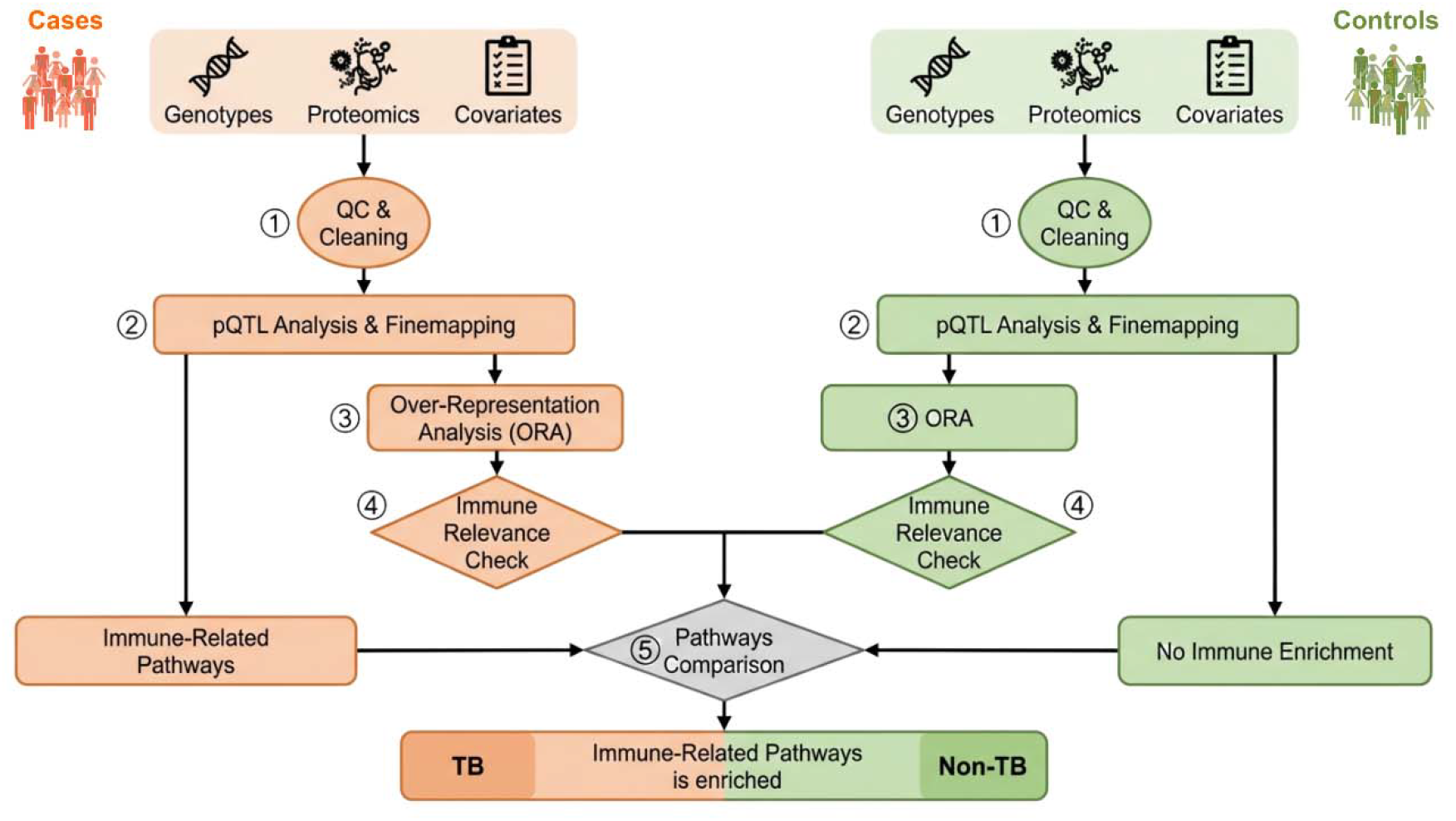
Overview of the pQTL analysis workflow. Separate cis-pQTL analyses were performed in TB progressors and matched controls using genotype, plasma proteomic, and covariate data. Significant pQTL-associated proteins were then evaluated by Gene Ontology over-representation analysis to compare pathway enrichment between groups. **ALT Text Figure 1:** Workflow diagram showing integration of plasma proteomics, genotype data, and clinical covariates including TB progression status in people with HIV. Separate cis-pQTL analyses were performed in TB progressors and matched non-progressing controls, followed by identification of significant pQTL-associated proteins and GO enrichment analysis.

Plasma proteomes were quantified by dia-PASEF mass spectrometry as described [8]. Genotyping used the Illumina Global Screening Array v3.0+MD or existing SHCS genetic data; quality control and imputation were performed per batch as described [9]. After quality-control filtering (variant call rate, Hardy-Weinberg equilibrium, minor allele frequency, and imputation quality thresholds applied per batch [9]), 5,531,557 variants remained in 60 cases and 194 controls. The first 10 genetic principal components were included as covariates to control population stratification.

We tested cis-variants against protein level using an additive linear model (MatrixEQTL v2.3 [10], TensorQTL v1.0.10 [11], GCTA v1.93.2 [12]), adjusting for age, sex, 10 genetic principal components, and proteomic batch. Independent variants were defined by LD clumping (r^2^<0.1, 1-Mb window) within each stratum; a pQTL was significant if it passed a protein-specific Bonferroni threshold (α=0.05/number of independent variants) and was concordant across all three tools. Fine-mapping used susieR (v0.14.2, default parameters except five rather than ten single effects [13]); we defined candidate variants as those with posterior inclusion probability (PIP) >0.9. TB-progressor and control analyses were conducted separately as exploratory screens; the resulting “progressor-associated” and “control-associated” labels are descriptive and do not, by themselves, constitute a formal test of between-group difference. To assess whether latent MTB infection shaped control-associated signals, we repeated the control analysis excluding participants with evidence of latent infection, using identical thresholds and tool-intersection criteria, and compared results descriptively.

For genes encoding significant pQTL-associated proteins, we performed over-representation analysis (clusterProfiler [14]) restricted to Gene Ontology biological process, cellular component, and molecular function terms, evaluated separately by stratum with Benjamini-Hochberg FDR correction.

## RESULTS

The pQTL analysis included 60 TB progressors and 194 matched controls (n=254). Proteomic profiling covered a median of 906 proteins per sample, with similar detection rates and sampling time points across groups [8].

Stratified analyses identified different cis-pQTL signals in progressors and controls (Figure 2). Progressors showed 26 significant cis-pQTL signals linked to 12 proteins, whereas controls showed 107 cis-pQTL signals linked to 14 proteins, with no protein shared between the two stratified sets. This non-overlap is exploratory and does not, by itself, establish differential genetic regulation between groups; it may partly reflect the roughly three-fold difference in control sample size.

**Figure 2.**
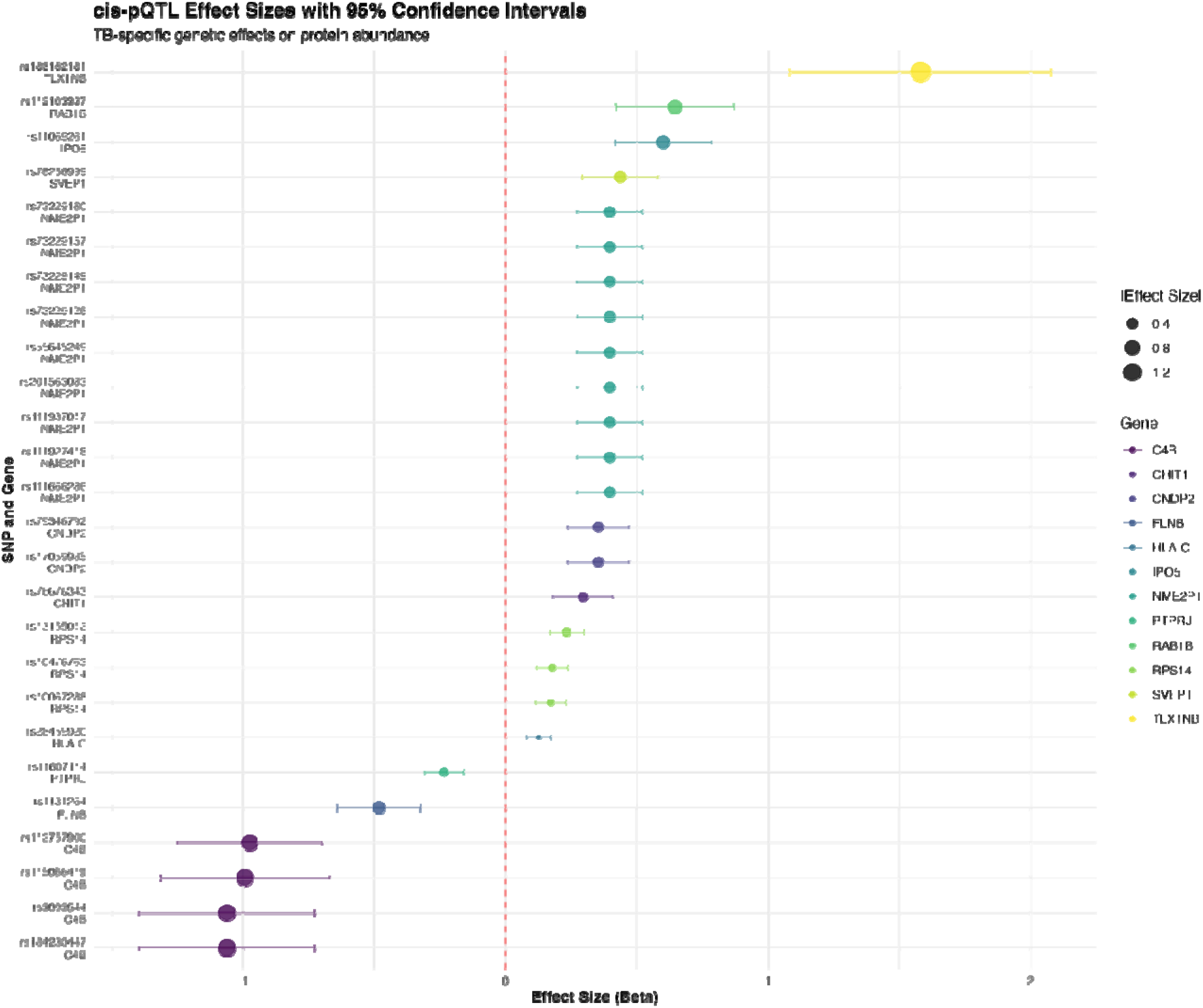
cis-pQTL signals in TB progressors. Forest plot of cis-pQTL effect sizes detected in the TB-progressor stratified analysis. Each point shows the estimated effect of a genetic variant on its corresponding protein level; horizontal lines show 95% confidence intervals. **ALT Text Figure 2:** Forest plot showing cis-pQTL effect estimates for proteins associated with TB progression in people with HIV. Points represent estimated genetic effects on plasma protein abundance, and horizontal lines represent confidence intervals.

Among progressor-associated signals, HLA-C was the most statistically robust (β=0.125, P<0.001); as the MHC class I molecule presenting pathogen-derived peptides to CD8+ T cells, even a modest genetically driven variation could plausibly relate to antigen presentation in TB. C4B, part of the classical complement pathway, showed the largest effect sizes among well-annotated genes (β range -1.061 to -0.975), linking the associated alleles to lower C4B levels; however, sensitivity analyses indicated this signal is unlikely to be exclusively progressor-specific and more likely reflects a difference in degree, rather than presence, of C4B genetic regulation between groups. CHIT1 (β=0.295), a macrophage-secreted chitinase proposed as a TB biomarker, and PTPRJ (β=-0.235), a protein involved in immune-cell signaling and phagocytosis-related pathways, were also detected among progressor-associated proteins. Additional signals with less direct functional annotation included IPO5, RAB1B, SVEP1, RPS14, CNDP2, and FLNB (β range -0.482 to 0.645); these may plausibly extend to vesicular trafficking, ribosomal, or metabolic processes relevant to activated immune cells, but this link is indirect and unconfirmed. TLX1NB showed the largest overall effect size (β=1.581), while NME2P1 also showed a positive association (β=0.396). Given the limited direct immune annotation of these loci, we treat both as signals of uncertain functional significance rather than candidate immune effector genes.

Control-associated pQTLs mapped to 14 proteins, including HLA-A, CFHR1, C7, and abundant plasma proteins such as ALB, APOM, and RARS1; these did not show a comparable immune-enrichment pattern in this analysis. Repeating the control analysis after excluding participants with latent MTB infection left most signals detectable, with a few attenuated and a few new signals emerging (Supplementary Table), suggesting the overall control pQTL architecture was largely, though not entirely, preserved.

Gene Ontology enrichment identified 46 immune-related biological processes among progressor-associated proteins (FDR-adjusted P<0.05) versus one in controls (“humoral immune response,” P=.0088; CFHR1, BST1, HLA-A, C7). Progressor-associated enrichment spanned MHC class I antigen presentation (P=.0133), lymphocyte-mediated immunity (P=.0011; C4B, SVEP1, HLA-C), classical complement activation (P=.0003) and opsonization (P=.0099), phagocytosis regulation (P=.0008) and Fcγ receptor signaling (P=.0151), and immune-regulatory processes such as negative regulation of humoral immune response (P=.0099), largely driven by SVEP1. These results describe a candidate immune-related module linking HLA-C, C4B, CHIT1, and PTPRJ in progressors; controls did not show a comparable pattern in this dataset.

## DISCUSSION

To our knowledge, this is the first pQTL analysis in PWH who progressed to active TB. Exploratory, stratified analyses identified a coherent set of genetically associated immune proteins in progressors that were not detected in non-progressors. This pattern is compatible with context-dependent genetic regulation, but stratified analyses alone do not formally test for a between-group difference; the absence of overlap may also partly reflect differences in statistical power between the two strata. Because germline variants precede disease, these signals are less likely to reflect purely reactive immune changes, although incipient subclinical disease cannot be excluded.

The progressor-associated proteins align with established TB immunology. HLA-C is a key molecule involved in antigen-presentation for CD8+ T-cell control of intracellular MTB, and HLA-region variants have long featured in TB genome-wide association studies [3,4]; our data identify a genetic association with pre-disease HLA-C abundance in progressors. C4B and complement activation are relevant to early opsonization of mycobacteria [15], and CHIT1 has been proposed as a macrophage activation marker [16]. PTPRJ points toward regulatory calibration of myeloid signaling alongside immune activation.

These findings are hypothesis-generating. Pending replication, formal interaction testing, and functional validation, they may help prioritize candidate proteins and pathways for future risk-stratification research; we do not consider the data sufficient to support specific therapeutic targeting or biomarker deployment at this stage.

This study has important limitations. The modest sample size (n=254) limits power for rare variants and trans-pQTLs. Although the cohort included participants from diverse ethnic backgrounds [8], generalizability to TB-endemic populations in Africa and Asia may still be limited by the Swiss cohort setting. The cross-sectional, single-timepoint design cannot capture dynamic changes in pQTLs, and the HIV-specific context may limit relevance to HIV-negative TB. We also lack an independent replication cohort, a formal genotype-by-group interaction test, and eQTL-pQTL colocalization; the small progressor-specific gene set makes pathway enrichment sensitive to individual genes, and residual, subclinical incipient TB cannot be fully excluded despite pre-diagnostic sampling.

Future work should prioritize replication in TB/HIV cohorts from high-burden settings, longitudinal pQTL profiling across disease stages, formal interaction testing, and eQTL-pQTL colocalization to clarify whether these genetically anchored proteins are mediators of TB progression or markers of an underlying process.

## Data Availability

The Swiss HIV Cohort Study (SHCS) established in 1988 collects clinical data and plasma samples from people living with HIV in Switzerland. All participants have provided written informed consent; however, strict confidentiality rules apply. Specifically, the SHCS is obligated to ensure that individuals cannot be re-identified from shared data, which prevents the release of detailed participant-level data (https://www.shcs.ch/for-researchers/open-data-statement-shcs/).

## ACKNOWLEDGMENTS

We thank all participants of the Swiss HIV Cohort Study (SHCS), and all study nurses, data managers, and physicians who contributed to data collection, as well as all members of the SHCS. Data were gathered from five Swiss university hospitals, two cantonal hospitals, 15 affiliated hospitals, and 36 private physicians (listed at https://www.shcs.ch/health-care-providers/).

The authors used AI-based tools (ChatGPT and Claude) to support language editing and improve the clarity of the manuscript. The authors reviewed and edited all AI-assisted content and take full responsibility for the final version of the article.

Author contributions: Conceptualization: JN, JF, IAA; Methodology: JN, JF, FJD, LB; Analysis: SB, MZ; Investigation: SB, MZ, JN, JF; Data curation: SB, MZ; Resources: CS, AC, MC, LE, PS; Writing – original draft: SB, MZ, CD, JN; Writing – review & editing: all authors; Supervision: JN, JF; Funding acquisition: JN.

## DATA AVAILABILITY

The SHCS collects clinical data and plasma samples from people with HIV in Switzerland under written informed consent and strict confidentiality rules that preclude release of participant-level data, consistent with the SHCS Open Data Statement (https://www.shcs.ch/for-researchers/open-data-statement-shcs/).

## REFERENCES

1. WHO. Global tuberculosis report 2025. Available from: https://iris.who.int/server/api/core/bitstreams/e97dd6f4-b567-4396-8680-717bac6869a9/content

2. Meintjes G, Maartens G. HIV-Associated Tuberculosis. N Engl J Med. 2024; 391(4):343–355.

3. Schurz H, Naranbhai V, Yates TA, et al. Multi-ancestry meta-analysis of host genetic susceptibility to tuberculosis identifies shared genetic architecture. eLife. 2024; 13:e84394.

4. Abel L, Fellay J, Haas DW, et al. Genetics of human susceptibility to active and latent tuberculosis: present knowledge and future perspectives. Lancet Infect Dis. 2018; 18(3):e64–e75.

5. Sun BB, Maranville JC, Peters JE, et al. Genomic atlas of the human plasma proteome. Nature. 2018; 558(7708):73–79.

6. Ferkingstad E, Sulem P, Atlason BA, et al. Large-scale integration of the plasma proteome with genetics and disease. Nat Genet. 2021; 53(12):1712–1721.

7. Scherrer AU, Traytel A, Braun DL, et al. Cohort Profile Update: The Swiss HIV Cohort Study (SHCS). Int J Epidemiology. 2021; 51(1):33–34j.

8. Kusejko K, Arefian M, Duroux D, et al. Inflammation and B cell activation define a plasma proteome signature predicting tuberculosis in people with HIV. mBio. 2025; 16(10):e0158525.

9. Schoepf IC, Thorball CW, Kovari H, et al. Polygenic Risk Scores for Prediction of Subclinical Coronary Artery Disease in Persons With Human Immunodeficiency Virus (HIV): The Swiss HIV Cohort Study. Clin Infect Dis. 2022; 76(1):48–56.

10. Shabalin AA. Matrix eQTL: ultra fast eQTL analysis via large matrix operations. Bioinformatics. 2012; 28(10):1353–1358.

11. Taylor-Weiner A, Aguet F, Haradhvala NJ, et al. Scaling computational genomics to millions of individuals with GPUs. Genome Biol. 2019; 20(1):228.

12. Jiang L, Zheng Z, Fang H, Yang J. A generalized linear mixed model association tool for biobank-scale data. Nat Genet. 2021; 53(11):1616–1621.

13. Wang G, Sarkar A, Carbonetto P, Stephens M. A simple new approach to variable selection in regression, with application to genetic fine mapping. J R Stat Soc: Ser B (Stat Methodol). 2020; 82(5):1273–1300.

14. Yu G, Wang L-G, Han Y, He Q-Y. clusterProfiler: an R Package for Comparing Biological Themes Among Gene Clusters. OMICS: A J Integr Biol. 2012; 16(5):284–287.

15. Mueller-Ortiz SL, Wanger AR, Norris SJ. Mycobacterial Protein HbhA Binds Human Complement Component C3. Infect Immun. 2001; 69(12):7501–7511.

16. Chen M, Deng J, Li W, et al. The relationship between chitotriosidase activity and tuberculosis. Epidemiology Infect. 2015; 143(15):3196–3202.

